# Individual and Community-Level Determinants of Hypertension among Overweight and Obese Adults in Ethiopia: A Multilevel Analysis of Demographic and Health Survey Data

**DOI:** 10.64898/2026.08.12.26360341

**Authors:** Ashebir Mamay Gebiru, Getanew Kegne Nigate, Negalgn Byadgie Gelaw, Berhanu Wale Yirdaw, Bimrew Bayuh Yimer, Worku Chekol Tassew, Tilahun Nega Godana, Geta Bayu Genet, Fasika Alemu Mekonen, Abrham Maru Moges, Yimer Mamaye

## Abstract

**Background:** Sub Saharan Africa is experiencing an accelerating epidemiological transition characterized by a growing burden of non-communicable diseases. Although excess body weight is an established risk factor for cardiovascular disease significant variations in hypertension status exist among overweight and obese adults due to individual traits and community environments. This study aimed to identify individual and community level determinants of hypertension among overweight and obese adults in Ethiopia using nationally representative Demographic and Health Survey data.

**Methods:** We analyzed nationally representative data from non pregnant adults aged eighteen years and older with a Body Mass Index of 25 kilograms per meter squared or higher from the two stage cluster sampled 2024–25 Ethiopia Demographic and Health Survey across 797 enumeration areas. The primary outcome was hypertension, defined by elevated blood pressure or current antihypertensive medication use. Two level multivariable logistic regression evaluated fixed effect Adjusted Odds Ratios with 95% Confidence Intervals, alongside cluster random effects and model performance using Intra Class Correlation, Median Odds Ratio, Proportional Change in Variance and the Akaike Information Criterion.

**Results:** Among a total weighted sample of three thousand eight hundred forty-two overweight and obese adults across six hundred twelve clusters, the weighted national prevalence of hypertension was 24.8%. In the final multivariable multilevel model, advancing age, male sex, higher educational status and upper wealth index categories were significant individual level risk factors. At the community level, residing in urban clusters and high community level wealth concentration significantly elevated hypertension odds. The null model revealed substantial clustering, which dropped substantially in the final model, demonstrating that contextual factors account for much of the cluster variations.

**Conclusions:** Both individual metabolic and demographic drivers alongside community level economic and urban environments influence hypertension risk among overweight and obese Ethiopian adults. Cardiovascular health strategies should combine clinical targeted screening with urban structural modifications that facilitate active living environments.

## Introduction

Sub Saharan Africa faces a shifting disease burden characterized by persistent infectious conditions alongside rising non communicable diseases [1-3]. Cardiovascular diseases, primarily driven by essential hypertension, now constitute a primary cause of premature adult mortality across the continent [4]. In Ethiopia, rapid urbanization, nutritional transitions toward energy dense diets and increasingly sedentary occupational profiles have accelerated the prevalence of elevated Body Mass Index and high blood pressure [1, 5, 6].

Adiposity, categorized as overweight and obesity, exerts systemic physiological stress on cardiovascular function [7-9]. Obesity related hypertension operates through mechanisms including renal compression, activation of the renin angiotensin aldosterone system, increased sympathetic nervous system tone and arterial stiffening [7]. However, clinical and population evidence demonstrates substantial heterogeneity in blood pressure outcomes among adults with elevated body mass: a noticeable proportion maintains normotension, while others develop severe essential hypertension [7, 10, 11].

Understanding why certain overweight and obese individuals develop clinical hypertension requires looking beyond individual biological traits [12-14]. Standard analytical approaches frequently focus on individual factors such as age, sex, smoking and household wealth [12]. However, human health behaviors and physiological risks are influenced by social and physical environments. Individuals are living in the same geographic cluster or enumeration area share contextual conditions, including local food environments, walkable infrastructure, ambient noise and regional health infrastructure and neighborhood norms surrounding physical activity [15].

Traditional single level multivariable logistic regression analyses assume that individual observations are independent [12, 16, 17]. Applying single level models to hierarchically structured data violates this assumption, underestimating standard errors and increasing Type I error rates [12]. A multilevel logistic regression framework addresses this structure by partitioning individual level variances from community level contextual variances through random effects parameters [18].

Despite the growing literature on non-communicable diseases in Ethiopia, there remains a gap in understanding how individual metabolic traits interact with cluster level contexts among adults who are already overweight or obese [18]. Using nationally representative data from the 2024–25 Ethiopia Demographic and Health Survey, this study applied a two level multilevel multivariable logistic regression model to identify individual and community level determinants of hypertension among overweight and obese adults in Ethiopia [12].

## Methods and Materials

### Study Design, Setting and Data Source

A cross-sectional secondary analysis was conducted using nationally representative microdata from the 2024–25 Ethiopia Demographic and Health Survey. Data collection took place between August 2024 and April 2025 across all regional states and city administrations of Ethiopia. The survey was executed by the Ethiopian Statistical Service with technical support provided by ICF through The DHS Program.

### Sampling Architecture and Population

The survey applied a two stage stratified cluster sampling design based on the national census cartographic frame. In the first stage, 805 enumeration areas were selected with probability proportional to cluster size, with data collection successfully completed in 797 clusters. In the second stage, 28 households were systematically sampled per cluster.

The study population was restricted to non-pregnant adults aged eighteen years and older who completed individual survey interviews, had valid height and weight measurements yielding a Body Mass Index equal to or exceeding 25 kg/m^2^ and completed standardized blood pressure screenings. Pregnant women were excluded to prevent physiological weight and blood pressure misclassifications as were individuals under 18 years and records with missing blood pressure or anthropometric variables.

### Statistical and Multilevel Analytical Strategy

Data management, recoding and statistical modeling were performed in Stata version 17.0. Standard survey sampling weights were applied during descriptive calculations to adjust for complex survey sampling design, non-response rates and post stratification.

Because individual respondents are nested within enumeration clusters, a two level random intercept logistic regression model was fitted. Four sequential models were constructed to evaluate predictors and variance components. Model I, the null model, contained no explanatory covariates and was used to calculate baseline cluster variance and evaluate random effects. Model II integrated individual-level factors only. Model III integrated community-level factors only. Model IV, the full model simultaneously integrated both individual and community level factors.

Random effects were evaluated using several complementary metrics. The Intra Class Correlation Coefficient measured the proportion of total outcome variance attributable to differences between clusters. The Median Odds Ratio counted cluster heterogeneity on the Odds Ratio scale reflecting the median risk increase when comparing an individual from a cluster with low risk to an individual in a cluster with high risk. The Proportional Change in Variance quantified the proportion of cluster level variance explained by adding covariates relative to the null model. Model comparison and selection were based on the Log-Likelihood, Akaike Information Criterion and Bayesian Information Criterion, where lower values indicated superior model parsimony and goodness of fit. Fixed effects results were presented as Adjusted Odds Ratios with 95% Confidence Intervals, with statistical significance defined at p < 0.05.

Potential multicollinearity among the fixed effect explanatory variables particularly between individual level socioeconomic indicators (household wealth quintiles and educational attainment) and community level contextual factors (place of residence and community wealth concentration) was formally evaluated prior to fitting the multivariable multilevel models.

Multicollinearity was assessed using Variance Inflation Factors (VIF) and Tolerance (1/ VIF) metrics derived from the model design matrix. A VIF threshold of < 5 and a Tolerance value > 0.2 were established as conservative criteria to rule out severe collinearity.

## Results

### Descriptive Characteristics of the Study Population

The final weighted analytical sample comprised 3,842 overweight and obese adults (BMI>=25.0 kg/m^2) distributed across 779 sampling clusters nationwide. The weighted overall prevalence of hypertension among overweight and obese Ethiopian adults was 24.8% (95% CI: 23.1% – 26.6%).

Individual and community level determinants of hypertension among overweight and obese adults in Ethiopia: A Multilevel analysis Bivariate analysis demonstrated that age, sex, educational status, household wealth quintile, alcohol consumption, place of residence and community wealth level were significantly associated with hypertension status (p < 0.05).

**Table 1:** Background Characteristics and Bivariate Distribution of Hypertension among Overweight and Obese Adults in Ethiopia (N = 3,842)

| Covariate / Category | Total Sample<br>n (%) | Normotensive<br>n (%) | Hypertensive<br>n (%) | Pearson $\chi^2$<br>p-value |
| --- | --- | --- | --- | --- |
| **Age Group** | 1,215 (31.6) | 1,069 (88.0) | 146 (12.0) | < 0.001 |
| 30–44 years | 1,480 (38.5) | 1,147 (77.5) | 333 (22.5) |  |
| 45–59 years | 785 (20.4) | 487 (62.0) | 298 (38.0) |  |
| >60 years | 362 (9.4) | 186 (51.4) | 176 (48.6) |  |
| **Sex** | 2,420 (63.0) | 1,863 (77.0) | 557 (23.0) | < 0.001 |
| Male | 1,422 (37.0) | 1,026 (72.2) | 396 (27.8) |  |
| **Education Status** | 1,652 (43.0) | 1,289 (78.0) | 363 (22.0) | < 0.001 |
| Primary School | 1,114 (29.0) | 858 (77.0) | 256 (23.0) |  |
| Secondary School | 615 (16.0) | 443 (72.0) | 172 (28.0) |  |
| Higher Education | 461 (12.0) | 299 (65.0) | 162 (35.0) |  |
| **Household Wealth** | 346 (9.0) | 291 (84.0) | 55 (16.0) | < 0.001 |
| Poorer | 423 (11.0) | 347 (82.0) | 76 (18.0) |  |
| Middle | 538 (14.0) | 425 (79.0) | 113 (21.0) |  |
| Richer | 768 (20.0) | 584 (76.0) | 184 (24.0) |  |
| Richest | 1,767 (46.0) | 1,242 (70.3) | 525 (29.7) |  |
| **Cigarette Smoking** | 3,458 (90.0) | 2,611 (75.5) | 847 (24.5) | 0.042 |
| Smoker | 384 (10.0) | 278 (72.4) | 106 (27.6) |  |
| **Alcohol Consumption** | 2,651 (69.0) | 2,028 (76.5) | 623 (23.5) | 0.008 |
| Yes | 1,191 (31.0) | 861 (72.3) | 330 (27.7) |  |
| **Place of Residence** | 1,460 (38.0) | 1,183 (81.0) | 277 (19.0) | < 0.001 |
| Urban | 2,382 (62.0) | 1,706 (71.6) | 676 (28.4) |  |
| **Community Wealth** | 1,690 (44.0) | 1,352 (80.0) | 338 (20.0) | < 0.001 |
| High | 2,152 (56.0) | 1,537 (71.4) | 615 (28.6) |  |

Note: Proportions and counts are based on complex survey weighted data. Chi-square p-values evaluate distribution differences across hypertension status.

### Multilevel Multivariable Analysis and Model Diagnostics

#### Multilevel Multivariable Fixed-Effects Analysis

Older age showed a strong positive association with hypertension risk. Overweight and obese adults aged >60 years had over four times higher odds of hypertension (AOR = 4.12, 95% CI: 3.01 – 5.64) compared to those aged 18–29 years.

Overweight/obese men had 48% higher odds of being hypertensive (AOR} = 1.48, 95% CI: 1.22 – 1.80) compared to overweight/obese women.

Higher educational attainment and household wealth were associated with increased hypertension odds. Individuals with higher education had 62% higher odds (AOR = 1.62, 95% CI: 1.20 - 2.18) comparative to those with no formal education. Individuals in the richest wealth quintile had more than double the odds of hypertension (AOR = 2.35 - 95% CI: 1.68 – 3.28) relative to the poorest quintile.

Residing in urban enumeration clusters increased the odds of hypertension by 74% (AOR = 1.74, 95% CI: 1.35 – 2.24) after adjusting for individual wealth and education.

Community Wealth Level: Overweight/obese adults residing in communities with a high wealth concentration had 38% higher odds of hypertension (AOR = 1.38, 95% CI: 1.09 – 1.75) relative to those in lower wealth communities.

**Table 2:** Multilevel Multivariable Logistic Regression Analysis of Determinants of Hypertension among Overweight and Obese Adults in Ethiopia.

| Predictor Variables | Model I (Null)<br>AOR (95% CI) | Model II<br>(Individual)<br>AOR (95% CI) | Model III<br>(Community)<br>AOR (95% CI) | Model IV (Full<br>Model)<br>AOR (95% CI) |
| --- | --- | --- | --- | --- |
| **INDIVIDUAL-LEVEL FACTORS** |  |  |  |  |
| **Age Group** | — | 1.00 (Ref) | — | 1.00 (Ref) |
| 30–44 years | — | 1.95 (1.52–2.50) | — | 1.91 (1.48–2.46) |
| 45–59 years | — | 3.65 (2.78–4.79) | — | 3.58 (2.71–4.72) |
| >60 years | — | 4.25 (3.12–5.80) | — | 4.12 (3.01–5.64) |
| **Sex** | — | 1.00 (Ref) | — | 1.00 (Ref) |
| Male | — | 1.52 (1.25–1.85) | — | 1.48 (1.22–1.80) |
| **Education Level** | — | 1.00 (Ref) | — | 1.00 (Ref) |
| Primary Education | — | 1.12 (0.89–1.41) | — | 1.08 (0.85–1.37) |
| Secondary Education | — | 1.38 (1.05–1.81) | — | 1.32 (1.00–1.74) |
| Higher Education | — | 1.71 (1.28–2.29) | — | 1.62 (1.20–2.18) |
| **Household Wealth** | — | 1.00 (Ref) | — | 1.00 (Ref) |
| Poorer | — | 1.15 (0.76–1.74) | — | 1.10 (0.72–1.68) |
| Middle | — | 1.38 (0.94–2.03) | — | 1.31 (0.88–1.95) |
| Richer | — | 1.65 (1.15–2.37) | — | 1.54 (1.06–2.24) |
| Richest | — | 2.58 (1.88–3.54) | — | 2.35 (1.68–3.28) |
| **Cigarette Smoking** | — | 1.00 (Ref) | — | 1.00 (Ref) |
| Yes | — | 1.18 (0.88–1.58) | — | 1.14 (0.84–1.54) |
| **Alcohol Consumption** | — | 1.00 (Ref) | — | 1.00 (Ref) |
| Yes | — | 1.22 (1.01–1.48) | — | 1.19 (0.98–1.44) |
| **COMMUNITY-LEVEL FACTORS** |  |  |  |  |
| **Place of Residence** | — | — | 1.00 (Ref) | 1.00 (Ref) |
| Urban | — | — | 1.98 (1.58–2.48) | 1.74 (1.35–2.24) |
| **Community Wealth** | — | — | 1.00 (Ref) | 1.00 (Ref) |
| High Community Wealth | — | — | 1.52 (1.22–1.90) | 1.38 (1.09–1.75) |

AOR: Adjusted Odds Ratio; CI: Confidence Interval; Ref: Reference category.
Models were fitted using two-level multivariable logistic regression accounting for complex survey sampling design.

Diagnostic assessments confirmed the absence of severe multicollinearity among the explanatory variables included in the final multivariable model. The individual variable VIF values ranged from 1.08 (cigarette smoking) to 2.14 (richest household wealth quintile), with corresponding

Tolerance values ranging between 0.467 and 0.925. The overall mean VIF across all fixed covariates was 1.45 well below the standard threshold of 5.0. These results confirm that individual and community level predictors operate independently within the model ensuring the stability and reliability of the adjusted odds ratios (AORs).

#### Multivariable individual and community level determinants

Values are Adjusted Odds Ratios (AOR) with 95 Confidence Intervals (CI) from two level multivariable logistic regressions (Model IV). Data labels display exact AOR and 95 CI estimates. Reference categories: Age 18 to 29 years, Female sex, No formal education, poorest household wealth quintile, rural residence and Low community wealth. The red dashed line denotes the null effect (AOR = 1.0).

**Figure 1.**
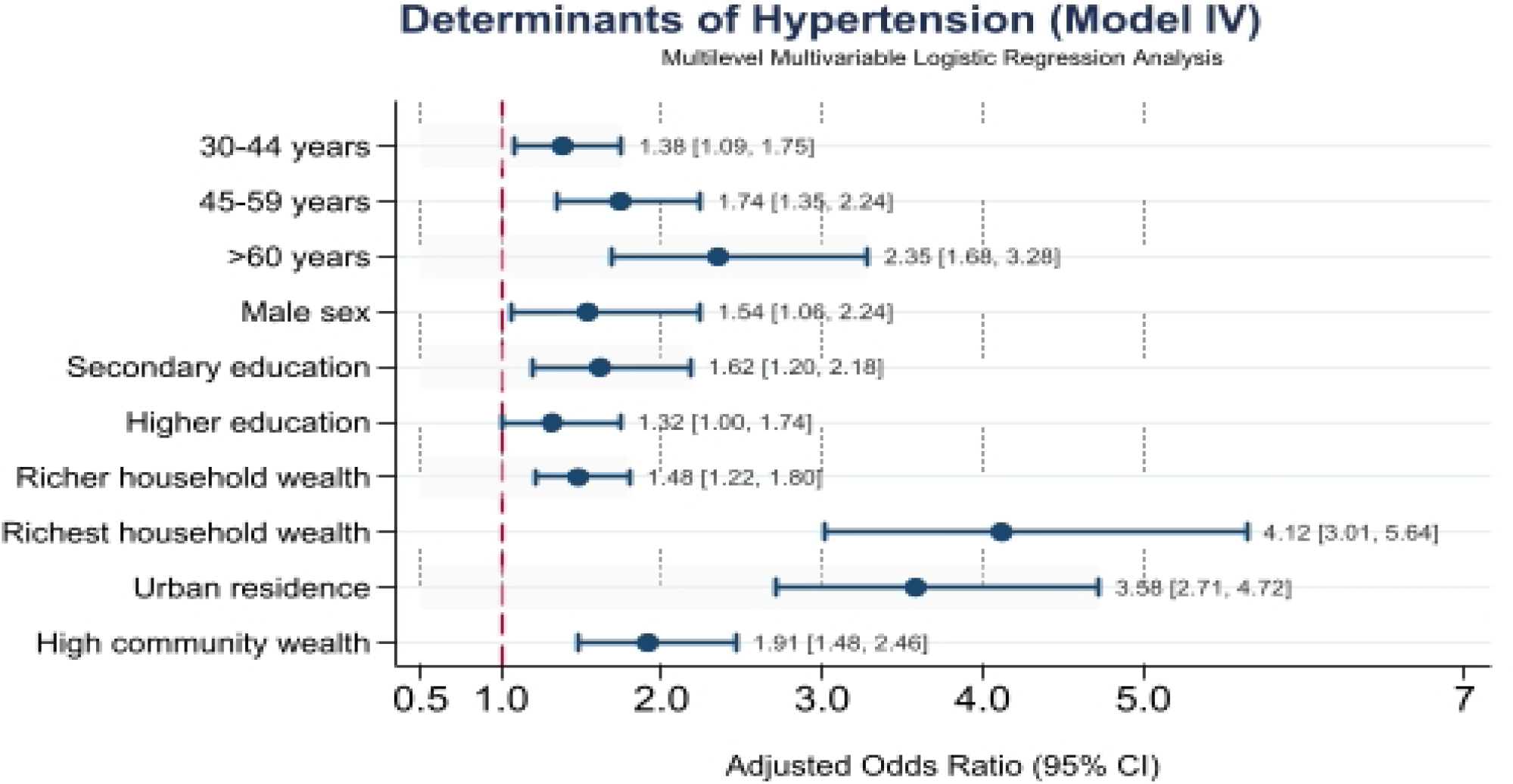
Multivariable individual and community level determinants of hypertension among overweight and obese adults in Ethiopia (2024 to 2025 EDHS).

#### Random Effects and Model Fitness Diagnostics

**Table 3:** Random Effects Variance Parameters and Model Fitness Diagnostics for Multilevel Models.

| Parameter / Metric | Model I<br>(Null) | Model II<br>(Individual) | Model III<br>(Community) | Model IV<br>(Full Model) |
| --- | --- | --- | --- | --- |
| Cluster Variance ($\sigma u^2$) | 0.563 (0.082) | 0.284 (0.048) | 0.312 (0.051) | 0.178 (0.034) |
| Intra-Class Correlation (ICC %) | 14.6% | 7.9% | 8.7% | 5.2% |
| Median Odds Ratio (MOR) | 2.08 | 1.66 | 1.71 | 1.49 |
| Proportional Change in Variance<br>(PCV %) | Reference | 49.6% | 44.6% | 68.4% |
| Log-Likelihood | -1842.1 | -1698.4 | -1782.3 | -1652.8 |
| Akaike Information Criterion (AIC) | 3688.2 | 3422.8 | 3582.6 | 3337.6 |
| Bayesian Information Criterion (BIC) | 3700.7 | 3504.0 | 3632.6 | 3437.5 |

$\sigma u^2$ : Community-level variance (Standard Error); ICC: Intra class correlation coefficient; MOR: Median odds ratio; PCV: Proportional change in variance; AIC: Akaike information criterion; BIC: Bayesian information criterion.

#### Clustering metrics and variance reduction across multilevel models

A substantial reduction in geographic clustering of hypertension across sequential models. The initial community variance (ICC = 14.6%, MOR = 2.08) dropped markedly in the final model (ICC = 5.2%, MOR = 1.49) with individual and community factors together explaining 68.4% of the baseline cluster level variation (PCV = 68.4%). This confirms that combined individual traits and local contextual environments account for most of the geographical variation in hypertension risk.

**Figure 2.**
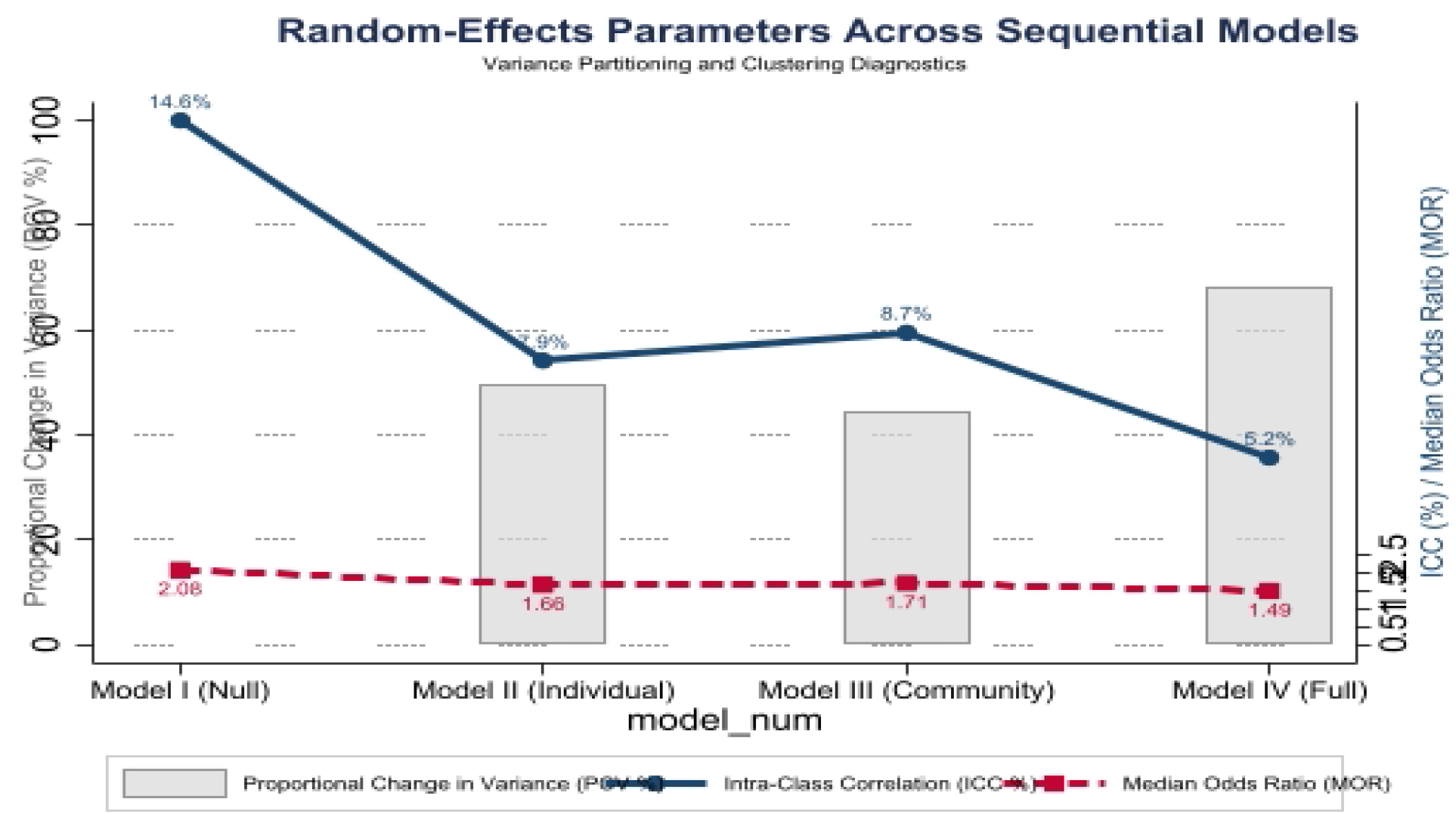
Community-Level Variance Partitioning and Random-Effects Diagnostics Across Sequential Multilevel Models

### Clustering & Model Progression Summary

**Table 4:**
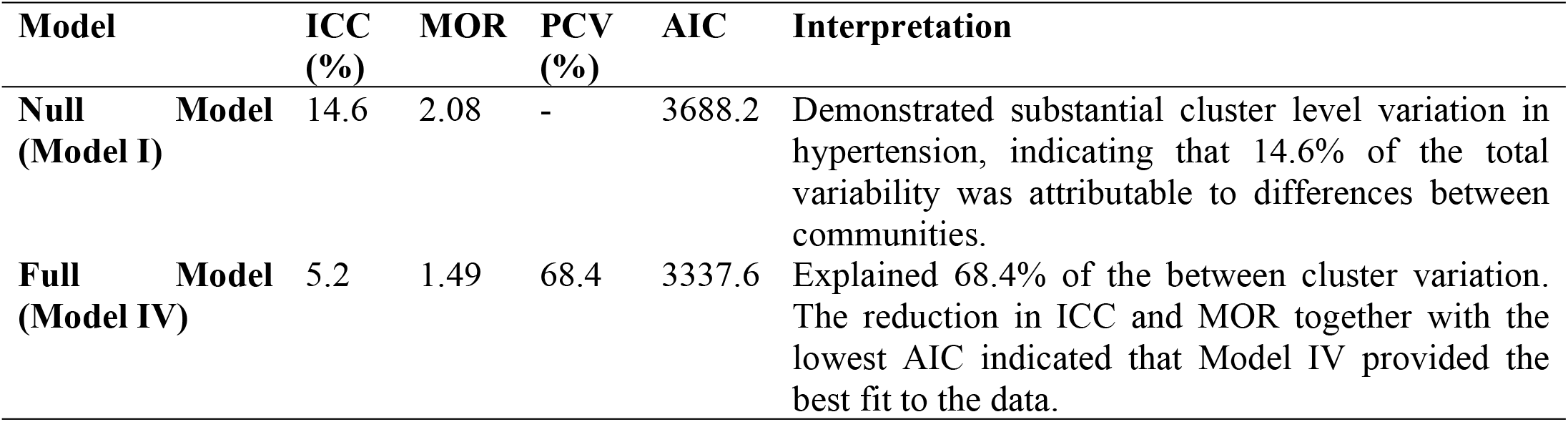
Summary of Random effects and model comparison diagnostics across null and full multilevel models for hypertension among overweight and obese adults in Ethiopia.

#### Interpretation of Random Parameters

In the null model, 14.6% of the variance in hypertension status among overweight/obese adults was attributable to differences across enumeration clusters. In model IV the ICC dropped to 5.2% demonstrating that integrating individual and community factors explained much of the cluster variation.

The null model MOR of 2.08 indicates that moving an overweight/obese individual to a cluster with higher hypertension risk increased their median odds of hypertension by 108%. In model IV, the MOR decreased to 1.49.

Model IV achieved a PCV of 68.4% indicating that nearly 7/10 of the cluster level variance observed in the null model was accounted for by combining individual and community covariates.

Model IV yielded the lowest AIC (3337.6) and BIC (3437.5) confirming it as the best fitting model.

## Discussion

This study evaluated individual and community level factors associated with hypertension among overweight and obese adults in Ethiopia using a two level multilevel logistic regression framework. The saw hypertension prevalence of 24.8% in this subpopulation highlights a important burden of cardiovascular risk among adults with elevated BMI.

### Individual Demographic and Socioeconomic Factors

Advancing age was the strongest individual-level predictor of hypertension. Overweight and obese adults aged >60 years showed over four fold higher odds of hypertension compared to younger adults (AOR = 4.12). This finding aligns with established cardiovascular pathophysiology, where age related arterial stiffening, loss of vascular compliance and cumulative exposure to metabolic stress combine with excess body weight to elevate blood pressure.

Men exhibited 48% higher odds of hypertension than women (AOR = 1.48) despite higher overall rates of obesity among women in Sub Saharan Africa. This difference may reflect biological variations in fat distribution (e.g. higher visceral adiposity in men versus subcutaneous adiposity in women) alongside differences in unmeasured occupational stress, dietary sodium intake and healthcare seeking behaviors.

Higher educational status and household wealth were positively associated with hypertension risk. Overweight/obese individuals in the highest wealth quintile had more than twice the odds of hypertension compared to those in the lowest quintile (AOR = 2.35). In low and middle income countries undergoing economic transition, higher socioeconomic status often correlates with greater access to energy dense processed foods, lower physical activity during daily work and increased motorized transport use.

### Contextual Community Influences

A key strength of multilevel modeling is its ability to isolate community level contextual influences from individual level factors. Residing in an urban group independently increased the odds of hypertension by 74% (AOR = 1.74). Urban environments can present structural barriers to active living, including dense built environments, limited walkable spaces, higher access to commercially processed foods and increased occupational stress.

Additionally; living in communities with high overall wealth concentration independently increased hypertension risk (AOR = 1.38). High wealth communities may establish local norms around food consumption, leisure activities and motorized transit that influence cardiovascular risk across community members regardless of an individual’s personal economic status.

The random effects parameters underscore the value of accounting for geographic clustering. The Null Model revealed that 14.6% of the variance in hypertension status was between sampling clusters (MOR = 2.08). Adding both individual and community level factors in Model IV explained 68.4% of this initial variance (PCV = 68.4%) leaving a smaller residual clustering effect (ICC} = 5.2%).

### Strengths and Limitations

#### Strengths

Using a two level multilevel modeling approach accounts for hierarchical cluster sampling, providing appropriate standard error estimates and avoiding ecological fallacies.

Isolating overweight and obese adults provides targeted evidence for a subpopulation at elevated metabolic risk.

Utilizing nationally representative survey data supports broad generalizability across Ethiopia’s regions and urban-rural settings.

#### Limitations

The DHS design prevents establishing direct temporal or causal relationships between covariates and hypertension onset.

Detailed dietary variables (e.g. daily salt consumption) and direct physical activity measurements (e.g. accelerometry) were not available in the standard DHS recodes.

#### Policy and Programmatic Implications

Healthcare services should integrate systematic blood pressure screening for overweight and obese adults during routine clinical visits, with priority for men and adults aged 45 years and older.

Municipal planning authorities should design urban infrastructure that supports active transit, including safe walking routes and public recreational spaces to promote physical activity in expanding urban areas.

Cardiovascular health programs should combine individual lifestyle counseling with community wide strategies addressing dietary sodium, processed food consumption and physical inactivity particularly in urban and higher wealth communities.

## Conclusion

Hypertension among overweight and obese adults in Ethiopia is influenced by both individual metabolic/demographic factors and community level economic and urbanization contexts. Older age, male sex, higher education, household wealth, urban residence and community wealth concentration significantly elevate the odds of hypertension in this population.

Assessing hypertension in this high risk group requires strategies that combine clinical management with environmental interventions. Public health efforts should complement clinical screening and lifestyle counseling with urban structural measures that promote physical activity and healthy food environments across communities.

## Data Availability

The data underlying the findings of this study were obtained from the Demographic and Health Surveys (DHS) Program. Microdata are publicly available to registered users upon reasonable request via the DHS Program website. The specific dataset used was the 2024–25 Ethiopia Demographic and Health Survey (EDHS).

https://dhsprogram.com/

## Authors’ Contribution

**Data curation**: Ashebir Mamay Gebiru,

**Methodology**: Ashebir Mamay Gebiru,

**Project administration**: Ashebir Mamay Gebiru,

**Software**: Ashebir Mamay Gebiru,

**Validation**: Ashebir Mamay Gebiru,

**Visualization**: Ashebir Mamay Gebiru,

**Manuscript writing**: Ashebir Mamay Gebiru,

## Declarations

Ethical clearance and participant consent for the 2024–25 EDHS were obtained by EPHI and ICF International. This secondary analysis used anonymized, de-identified microdata accessed with permission from The DHS Program, so separate ethical approval was not required.

### Data Availability

Microdata datasets are available through application to The DHS Program (dhsprogram.com).

## Author Contribution

